# Relationship between Striatal Connectivity and Apathy during Phosphodiesterase 10 Inhibition in Schizophrenia

**DOI:** 10.1101/2024.04.13.24305575

**Authors:** Wolfgang Omlor, Giacomo Cecere, Gao-Yang Huang, Finn Rabe, Nils Kallen, Matthias Kirschner, Werner Surbeck, Achim Burrer, Tobias Spiller, George Garibaldi, Štefan Holiga, Juergen Dukart, Daniel Umbricht, Philipp Homan

## Abstract

Negative symptoms in schizophrenia remain a challenge with limited therapeutic strategies. The novel compound RG7203 promotes reward learning via dopamine D1-dependent signaling and therefore holds promise to improve especially the apathy dimension of negative symptoms. When tested as add-on to antipsychotic medication apathy did not change significantly with RG7203 versus placebo. However, the response varied across patients, and a subset showed clinically relevant improvement of apathy. It remains unclear if these interindividual differences are related to neurobiological correlates. Due to the predominant binding of RG7203 in the striatum, we asked how apathy changes with RG7203 are related to changes in cortico-striatal connectivity. We focused on cortico-striatal circuits that have been associated with apathy and previously showed connectivity alterations in schizophrenia. In a double-blind, 3-way randomized crossover study, resting state functional magnetic resonance imaging was acquired in 24 individuals with schizophrenia following a 3-week administration of placebo, 5mg or 15mg of RG7203 as add-on to antipsychotics. We found that 5mg or 15mg of RG7203 did not lead to significant changes in striatal connectivity. However, changes in the apathy response across individuals were reflected by striatal connectivity changes. Apathy improvement with 5mg RG7203 vs. placebo was associated with increased connectivity between ventral caudate (vCaud) and paracingulate gyrus (PCG) as well as anterior cingulate cortex (ACC). The same trend was observed for 15mg RG7203 vs. placebo. Importantly, such associations were not observed for the negative symptom dimension of expressive deficits. These findings suggest that the relationship between vCaud-PCG/ACC connectivity and apathy response with RG7203 should be further explored in larger clinical studies. Replication and further elaboration of these findings could help to advance biologically informed treatment options for negative symptoms.

## Introduction

Negative symptoms in schizophrenia are a therapeutic challenge [1, 2]. While traditional antipsychotics treat positive symptoms, their effectiveness against negative symptoms is limited [1, 3]. This creates a crucial gap in treatment options, as negative symptoms are considered one of the main drivers of impaired functional outcomes and reduced quality of life [4].

Evidence from factorial analyses of different psychometric scales suggests that negative symptoms can be mapped onto the two dimensions apathy and diminished expression [5-7]. While the diminished expression dimension includes the domains blunted affect and alogia, the apathy dimension subsumes the domains avolition, asociality and anhedonia [8]. Individuals with schizophrenia show impaired learning from rewards which is mediated by dopamine-dependent D1-receptor signaling in the striatum [9] and related to motivation. Growing evidence suggests that these motivational and reward-related mechanisms preferentially underlie the apathy dimension of negative symptoms [8, 10-15]. The recent Phosphodiesterase 10 inhibitor RG7203 mainly binds in the striatum to enhance D1- and dampen D2-signaling. The D1-dependent pathway is important for positive reward learning, while the D2-dependent pathway has been implicated in learning from negative reward [9]. By enhancement of D1-signaling, RG7203 promotes reward learning and therefore holds promise to improve particularly the apathy dimension of negative symptoms [16]. While RG7203 has shown small, but consistent positive effects across four different paradigms probing reward functioning in healthy volunteers [17], an add-on three-way, placebo-controlled cross-over study in schizophrenia patients with negative symptoms and stable antipsychotic treatment was negative overall [16]. While a subset of subjects showed clinically relevant apathy improvement, there were also patients with clinically relevant worsening of apathy. Instead of random longitudinal apathy fluctuations these interindividual differences could reflect a heterogeneous effect of RG7203 across patients. It is conceivable that patient-specific D1/D2-signaling features and differing D2-blockade of background antipsychotics could have led to a deleterious effect in certain patients due to the additional D2 blockade by RG7203, while the beneficial effect of enhanced D1-signaling may have prevailed in others [16]. Striatal functional connectivity during rest, a neurobiological indicator of synchronized neural activity in intra-striatal and cortico-striatal circuits, is related to striatal D1- and D2-signaling as well as to negative symptoms of schizophrenia [18-26].

We therefore asked if different apathy changes with RG7203, for example due to patient-specific D1/D2-signaling features such as the D1/D2 receptor ratio[26], are related to changes in cortico-striatal connectivity. We expected that different apathy changes with RG7203 will be related to connectivity changes in those cortico-striatal circuits that have been associated with apathy [8] and showed reduced connectivity in schizophrenia [23]. Specifically, we hypothesized that apathy improvement with RG7203 versus placebo will be associated with increased striatal connectivity and vice versa.

## Methods

### Participant Criteria and Experimental Design

Patients aged between 15 and 50 years with a confirmed DSM-5 diagnosis of schizophrenia were enrolled. Participants were required to have a minimum score of 18 on the PANSS negative symptom factor score [27] during the initial screening and to be in a symptomatically stable state. Participants on antipsychotic treatment were included if the dosage did not surpass the equivalent of 6 mg risperidone. Other inclusion criteria included: a score of 3 or above on the Clinical Global Impression Severity scale (indicating at least mildly ill); a PANSS depression score (G6) of 4 or below (indicating moderate or mild symptoms); and a score of 8 or below on the Calgary Depression Rating Scale for Schizophrenia. Exclusion criteria encompassed a score exceeding 2 (indicative of mild conditions) on any of the Clinical Global Impression Severity scale items of the Extrapyramidal Symptom Rating Scale and treatment with either olanzapine or clozapine within the preceding 3 months. An exhaustive list of eligibility criteria can be found in the Supplementary Material.

This study was a three-way, placebo-controlled cross-over study. Individuals with schizophrenia underwent randomization to one of six treatment sequences that were fully counterbalanced and characterized by three treatment periods with placebo, 5mg or 15mg of RG7203 (**Fig. 1a**, daily applied in identical oral capsules). Each sequence had about 8 participants and RG7203 was given as add-on treatment to stable antipsychotic background medication [16]. During the period of 15mg treatment, dosing was progressively increased to the set target over the initial week. A single treatment phase spanned three weeks, succeeded by a two-week wash-out period. fMRI scans and behavioral tasks were conducted on the concluding day of each treatment phase (day 22, **Fig. 1a**). Weekly evaluations were conducted to determine safety, compatibility, and psychological condition. A subsequent follow-up was performed approximately two weeks post the final medication dose. Adherence to the regimen was tracked using a mobile application.

**Figure 1:**
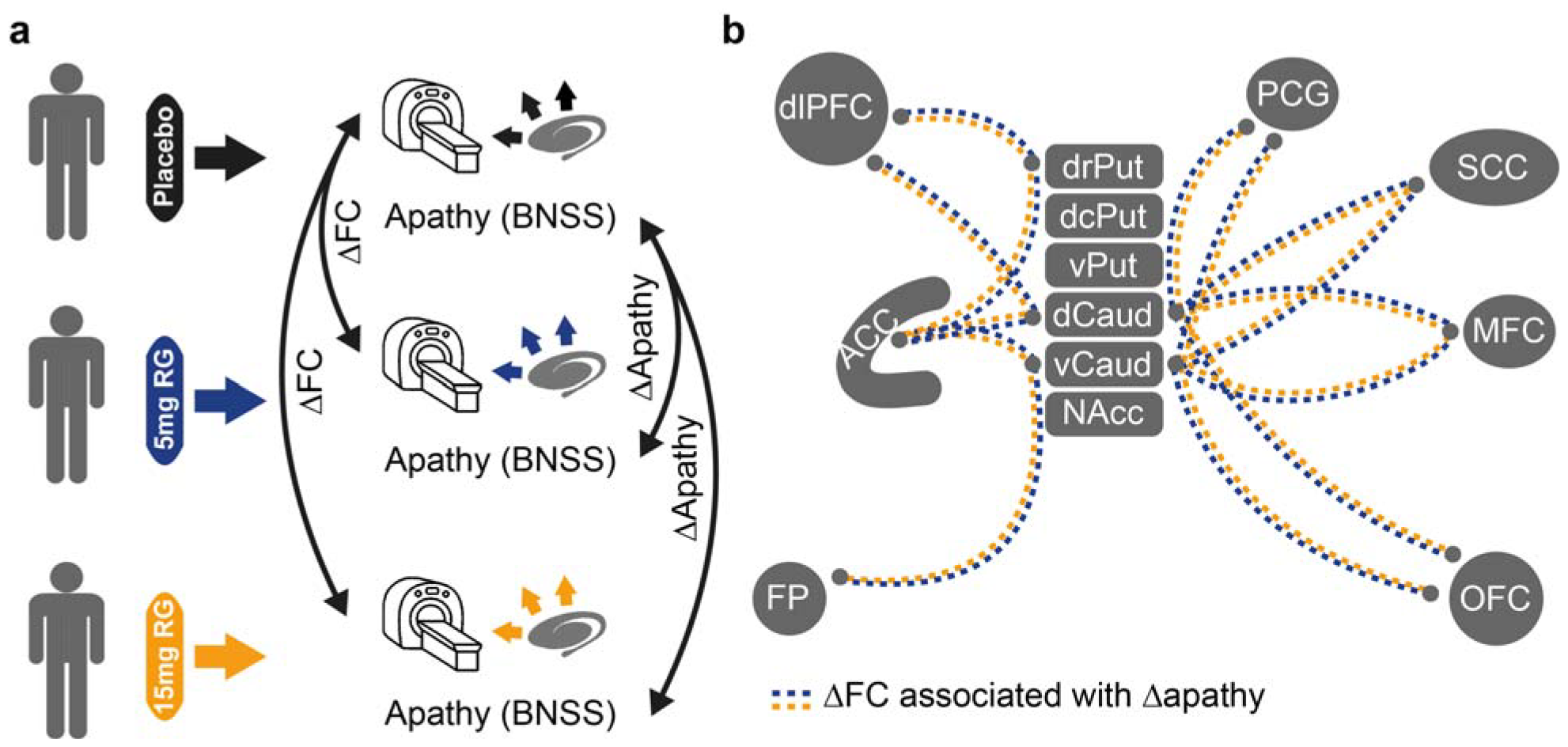
Hypotheses about effects of RG7203. **a**. Placebo, 5mg or 15mg RG7203 are given to individuals with schizophrenia, in addition to antipsychotic background medication. After placebo, 5mg or 15mg RG7203 have been given for three weeks, individuals with schizophrenia underwent fMRI and BNSS testing. Apathy scores were computed from the BNSS subdimensions avolition, asociality and anhedonia. ΔApathy was defined as the treatment effect on apathy and computed for 5mg RG7203 (5mg RG7203 minus placebo) and 15mg RG7203 (15mg RG7203 minus placebo). The treatment effect on cortico-striatal functional connectivity was defined as ΔFC and computed for 5mg RG7203 (5mg RG7203 minus placebo) and 15mg RG7203 (15mg RG7203 minus placebo). **b**. Cortico-striatal circuitry that has been associated with apathy [8] and that showed altered functional connectivity in individuals with schizophrenia and first-episode psychosis across previous studies [23]. We hypothesized that ΔFC in this circuitry correlates with Δapathy at 5mg (blue dotted line) and 15mg RG7203 (orange dotted line). Abbreviations: dr: dorsorostral; dc: dorsocaudal; v: ventral; d: dorsal; Put: Putamen; Caud: Caudate; NAcc: Nucleus accumbens; FP: Frontal pole; dlPFC: dorsolateral prefrontal cortex. ACC: Anterior cingulate cortex; PCG: Paracingulate gyrus; SCC: Subcallosal cortex; MFC: Medial frontal cortex; OFC: Orbitofrontal cortex.

Cognitive functions, such as reward-driven effortful behavior, probabilistic learning, and working memory, were assessed via fMRI (employing MID and n-back) in addition to behavioral tasks (the working memory reinforcement learning task [WMRLT] and effort-cost-benefit task [ECBT]). Results of these behavior tasks were presented previously [16]. Symptomatology was assessed with the PANSS, BNSS and CGI. The negative symptom dimensions apathy and diminished expression were derived from the BNSS [28]. The research protocol was registered on ClinicalTrials.gov (NCT02824055) and obtained approvals from responsible ethics oversight bodies (Copernicus Group IRB, P.O. Box 110605, Research Triangle Park, NC 27709, approval given on May 27^th^ 2016; Washington University in St. Louis, Human Protection Office, 660 South Euclid Ave., Campus Box 8089, St. Louis, MO 63110, approval given on 5^th^ August 2016; Alpha IRB, 1001 Avenida Pico, Suite C#497, San Clemente, CA 92673, approval given on 8^th^ July 2016; Integ Review IRB, 3815 S. Capital of Texas Hwy, Suite 320, Austin, TX 78704, approval given on 27^th^ May 2016).

### MRI Data Collection

We used three different 3T scanner models (GE 3T Discover 750w 25.0 by GE Healthcare; Siemens 3T MAGNETOM Trio and Siemens 3T Verio by Siemens Healthineers). Across all locations, BOLD fMRI information was gathered using a T2-weighted echo-planar imaging protocol (with parameters: repetition time of 2000 ms, echo time of 27 ms, flip angle set at 90°, 39 slices, and a voxel dimension of 3 × 3 × 3 mm with a 1 mm gap). For each patient, a conventional structural T1-weighted scan (1 × 1 × 1 mm) was also captured for alignment aims using default sequences at every imaging facility.

### fMRI Data Processing

The fMRI datasets underwent preprocessing via the Conn-Toolbox and were also analyzed using MATLAB (R2023a version by The MathWorks, Inc.). Preprocessing included realignment for motion and distortion correction, co-registration to structural scans, followed by standardization to the Montreal Neurological Institute coordinates, masking of non-gray matter voxels, and application of a 6mm full width at half maximum Gaussian smoothing kernel. We finally conducted an analysis of functional connectivity during rest using the Conn toolbox in Matlab (MathWorks, version R2023a).

### Statistical analyses and hypotheses

In a hypothesis-driven approach, we investigated the effect of RG7203 vs. placebo on functional connectivity in preselected components of the cortico-striatal circuit. Cortico-striatal components were selected if they have been implicated in apathy [8] and showed consistent alteration of functional connectivity in prior studies in schizophrenia and first-episode psychosis [23]. Apathy has been associated with the following cortico-striatal components: Anterior cingulate cortex (ACC), orbitofrontal cortex (OFC), ventromedial prefrontal cortex, dorsolateral prefrontal cortex (dlPFC) as well as the ventral and dorsal striatum [8]. Our definition of the ventromedial prefrontal cortex thereby included frontal pole (FP), medial frontal cortex (MFC), subcallosal cortex (SCC), paracingulate gyrus (PCG), ACC and OFC [29]. The striatum was subdivided into ventral rostral putamen (vPut), dorsal rostral putamen (drPut), dorsal caudal putamen (dcPut), ventral caudate (vCaud) and nucleus accumbens (NAcc) according to previously established coordinates [18]. The criteria above resulted in 14 connectivities between cortico-striatal component pairs that have been implicated in apathy and showed consistent connectivity alteration in schizophrenia and first-episode psychosis (**Fig. 1b**). From the 14 connectivities (**Fig. 1b**), 13 showed reduced connectivity in prior studies in schizophrenia and first-episode psychosis; only the connectivity between vCaud and OFC was reported to be increased [23]. Our primary hypothesis was that that improvement of apathy during treatment with RG7203 would be associated with increased connectivity in all 13 component pairs with reduced connectivity (and vice versa). In contrast, we expected that improvement of apathy during treatment with RG7203 would be associated with decreased connectivity between vCaud and OFC as this connectivity has been reported to be increased in schizophrenia [23]. Furthermore, we expected that similar correlations would not be expected for the dimension of diminished expression.

Changes in connectivity and apathy with 5mg or 15mg RG7203 vs. placebo were tested using a Wilcoxon signed-rank test. Correlations between changes in apathy and connectivity with 5mg or 15mg RG7203 vs. placebo were computed using the Spearman correlation coefficient. The 95% confidence intervals of the correlations and p-values of the correlations were computed. For all analyses with multiple comparisons, p-values were corrected according to the Benjamini-Hochberg procedure as implemented in the Matlab mafdr-function.

## Results

Across subjects, apathy did not change significantly for 5mg or 15mg of RG7203 vs. placebo (**Fig. 2a**). In the context of the BNSS scale from 0 (absence of symptoms) to 6 (severe symptoms), apathy changes spanned a clinically relevant range of 3.43 (SD: 0.84) and 3.71 (SD: 0.9) for 5mg and 15mg of RG7203 vs. placebo, respectively (**Fig. 2a)**. At both doses of RG7203 vs. placebo, 59% of the patients changed more than 0.5 points on the BNSS-scale (5mg RG7203: 21% with improvement; 15mg of RG7203: 25% with improvement). The apathy changes at 5mg and 15mg of RG7203 were positively correlated (r = 0.66; p = 4.07*10^−4^, **Fig. 2b**). We would expect that apathy changes at 5mg and 15mg of RG7203 are uncorrelated if they simply reflected spontaneous longitudinal apathy fluctuations. Due to the interindividual differences of apathy changes and their similarity at both doses of RG7203, we further explored their potential relationship to cortico-striatal connectivity.

**Figure 2:**
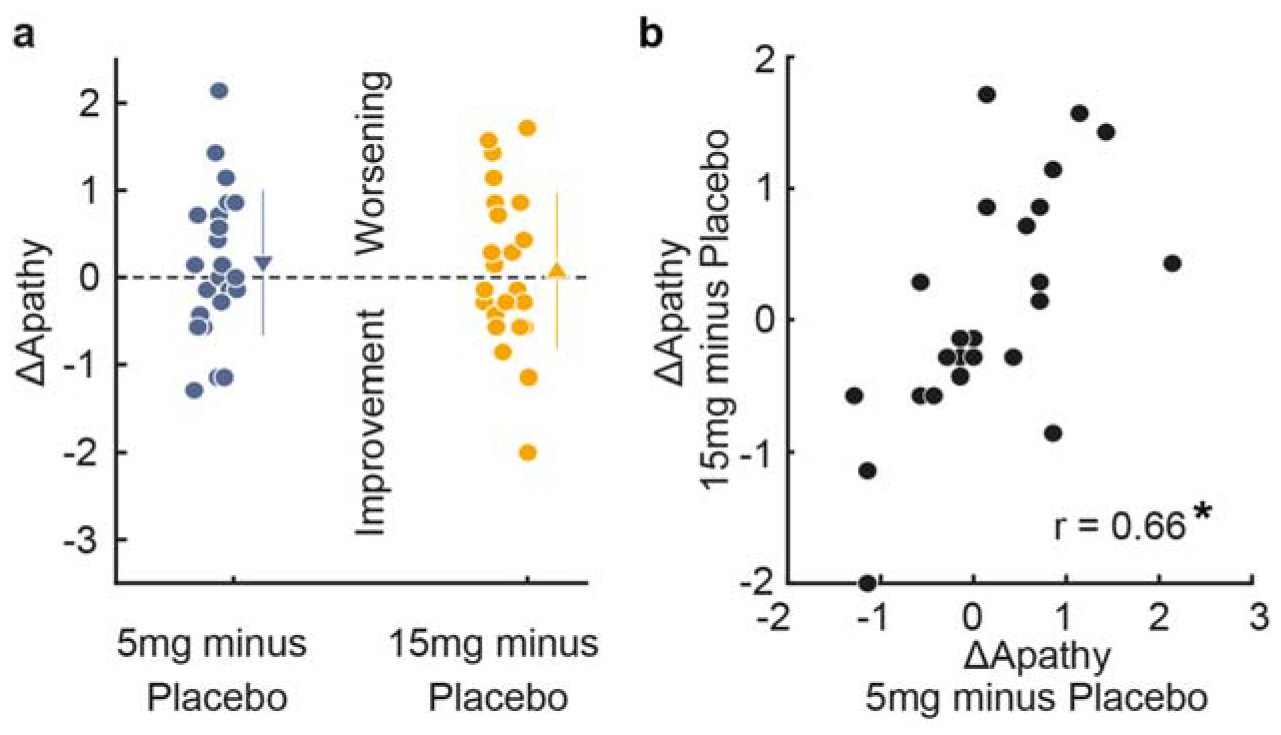
Apathy changes with RG7203 vs. placebo. **a**. Apathy changes (ΔApathy) when 5mg or 15mg RG7203 are given instead of placebo (5mg minus placebo and 15mg minus placebo); filled circles represent individual subjects; triangles with error bars show means and standard deviations. **b**. Correlation between ΔApathy at 5mg and 15mg RG7203. Asterisk indicates p<0.05.

In the apathy-related corticostriatal circuit, striatal connectivity did not change significantly for 5mg or 15mg RG7203 vs. placebo (**Fig. 3a, b**). At 5mg and 15mg RG7203, the highest interindividual differences were found for connectivity changes between vCaud and PCG (Range: 1.58 and 1.24; SD: 0.31 and 0.26, respectively), and the lowest interindividual differences were found for connectivity changes between dCaud and PCG (Range: 0.83 and 0.64; SD: 0.16 and 0.12, respectively). Striatal connectivity changes at 5mg and 15mg RG7203 were positively correlated for all circuit connections (p = 1.22*10^−4^, Wilcoxon signed-rank test for n = 14 correlation values, **Supplementary Fig. 1**), and the highest correlations were found for the connectivity pairs vCaud-PCG (r = 0.72, p = 1*10^−3^) and vCaud-ACC (r = 0.64, p = 6*10^−3^). We expected that connectivity changes at 5 and 15mg of RG7203 are uncorrelated if they simply reflect longitudinal fluctuations of functional connectivity. Due to the interindividual differences of connectivity changes and their similarity at both doses of RG7203, we then investigated if apathy changes with 5mg of RG7203 versus placebo (**Fig. 2a**) are reflected by corresponding changes in striatal connectivity (**Fig. 3a, b**): There was a negative correlation with connectivity changes between vCaud-PCG (r = -0.58, p = 0.02) and vCaud-ACC (r = -0.57, p = 0.02, **Fig. 3c, Supplementary Fig. 2a, b**). For 15mg of RG7203, we also found a negative correlation between changes in apathy and vCaud-PCG/ACC connectivity, but this relationship was not significant after FDR-correction (**Fig. 3c**, vCaud-PCG: r = -0.51, p = 0.09; unadjusted p = 0.01; vCaud-ACC: r = -0.5, p = 0.09; unadjusted p = 0.01). In summary, improvement in apathy during treatment with 5 mg of RG7203 was correlated with increased vCaud-PCG/ACC connectivity and vice versa.

**Figure 3:**
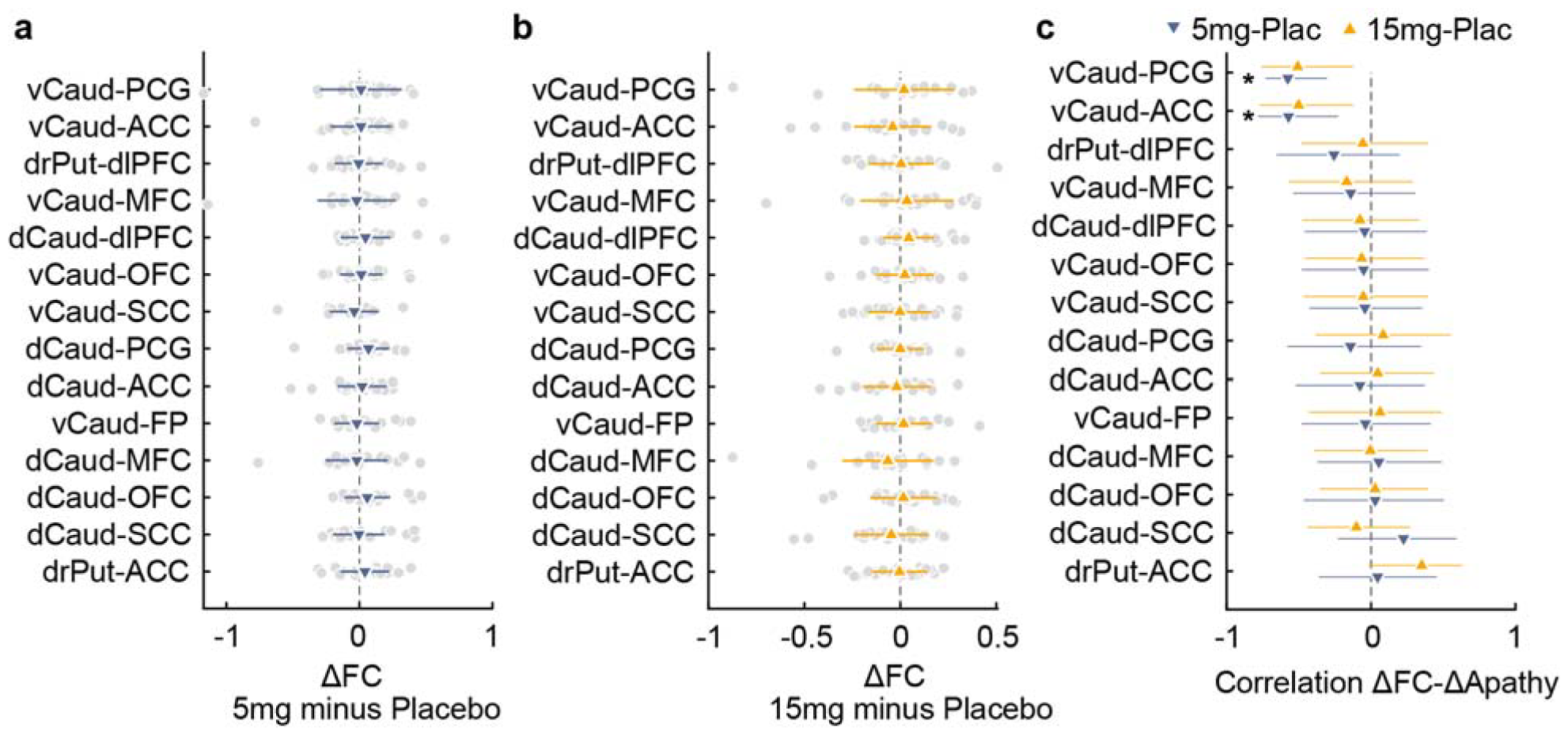
Correlation between changes in apathy and functional connectivity at 5mg and 15mg of RG7203. **a**. Functional connectivity changes (ΔFC) in the cortico-striatal circuit when 5mg RG7203 are given instead of placebo; filled circles (gray) represent individual subjects; blue downward-pointing triangles with error bars show mean and standard deviation. **b**. Functional connectivity changes (ΔFC) in the cortico-striatal circuit when 15mg RG7203 are given instead of placebo; filled circles (gray) represent individual subjects; orange upward-pointing triangles with error bars show mean and standard deviation **c**. Correlation between ΔApathy (apathy changes with 5mg or 15mg RG7203 vs. placebo) and ΔFC is illustrated for the cortico-striatal circuit. The error bars correspond to the 95% confidence interval. Asterisks indicate p<0.05.

We next computed connectivity alterations for all possible combinations of cerebrum structures at 5mg and 15mg RG7203 and explored their correlation with changes in apathy (**Fig. 4a, b**). This resulted in 1770 unique correlations (i.e. only the left lower triangle without the diagonal part from **Fig. 4a, b**, left panel). As a further test of the significance of our findings we compared the correlation between changes in apathy and vCaud-PCG/ACC connectivity with all possible correlations of changes in apathy and connectivity between all cerebrum structures. For this analysis, only the absolute value of correlations was used (**Fig 4a, b**, right panel). The correlation between changes in apathy and vCaud-PCG/ACC connectivity was above the 99^th^ percentile of all possible correlations for 5mg RG7203 vs. placebo, and around the 99^th^ percentile for 15mg RG7203 vs. placebo (**Fig. 4a, b**, vCaud-PCG: 5mg minus Placebo: 99.4th percentile; 15mg minus Placebo: 99th percentile; vCaud-ACC: 5mg minus Placebo: 99.4^th^ percentile; 15mg minus Placebo: 98.9^th^ percentile).

**Figure 4:**
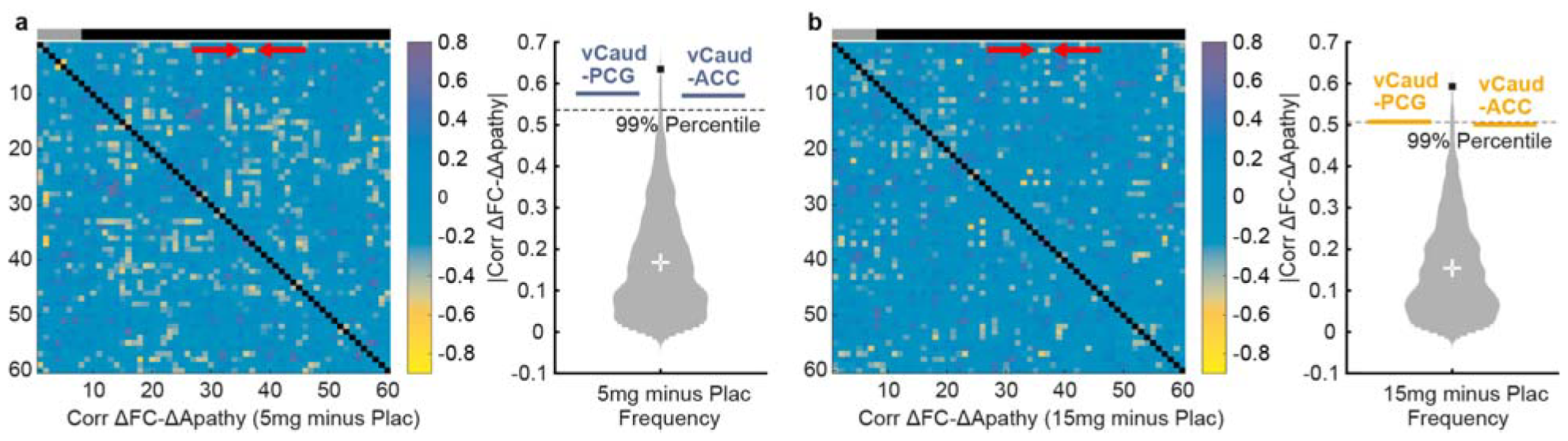
Relationship between changes in apathy and connectivity with RG7203 vs. placebo for all combinations of cerebrum structures. **a**. 5mg RG7203 compared to placebo. <u>Left panel:</u> Functional connectivity changes at 5mg RG7203 (ΔFC) has been computed for all possible combinations of the 60 anatomical cerebrum structures and correlated with corresponding apathy changes (ΔApathy); gray bar indicates striatal regions, black bar extrastriatal regions; correlation of ΔApathy and ΔFC between vCaud and PCG/ACC is indicated by red arrows. <u>Right panel:</u> Violin plot showing the absolute value of all 1770 unique correlations between ΔFC and Δapathy; correlation between Δapathy and ΔFC for the connectivity vCaud-PCG and the connectivity vCaud-ACC is indicated by blue bars. The black square indicates the maximum of all correlations. **b**. 15mg RG7203 compared to placebo: Same computations and conventions as in a, but correlation between ΔApathy and ΔFC for vCaud-PCG/ACC is indicated by orange instead of blue bars in the right panel.

To further contextualize our findings, we then added several explorative analyses. First, connectivity changes with 5mg or 15mg RG7203 versus placebo did not correlate with changes in diminished expression (**Supplementary Fig. 3a, b**). We then investigated if striatal connectivity under placebo (**Supplementary Fig. 4a**) is related to apathy changes when 5mg or 15mg RG7203 are given versus placebo. As increased vCaud-PCG/ACC connectivity with 5mg of RG7203 was a correlate of better apathy response, we hypothesized that better apathy response at RG7203 would also be associated with lower and more pathological [23] vCaud-PCG/ACC connectivity under placebo. Connectivity between vCaud and PCG as well as between vCaud and ACC during placebo treatment was positively correlated with apathy changes at 5mg and 15mg RG7203 (vCaud-PCG: 5mg minus placebo, r = 0.67; p = 4*10^−3^; 15mg minus Placebo: r = 0.72; p = 5*10^−4^; vCaud-ACC: 5mg minus placebo: r = 0.61; p = 1*10^−2^; 15mg minus placebo: r = 0.75; p = 4*10^−4^, n = 24; **Supplementary Fig. 4b, Supplementary Fig. 5a, b**). Thus, lower vCaud-PCG/ACC connectivity under placebo was associated with a better apathy response at 5mg and 15mg RG7203 vs. placebo. With regard to the correlation with apathy alterations at RG7203, we also compared vCaud-PCG/ACC connectivity under placebo with connectivity of all other cerebrum structures under placebo. The correlations between apathy changes and vCaud-PCG/ACC connectivity under placebo were well above 99^th^ percentile of all possible correlations, both at 5mg and 15mg of RG7203, or even represented the maximal correlation (**Supplementary Fig. 6a, b**, right panel; vCaud-PCG: 5mg minus Placebo: 99.9th percentile and second largest value; 15mg minus Placebo: 99.9th percentile and second largest value; vCaud-ACC: 5mg minus Placebo: 99.8^th^ percentile and fifth largest value; 15mg minus Placebo: Maximal correlation). Thus, the correlation magnitudes for the connections vCaud-PCG and vCaud-ACC were the two greatest at 15mg and among the top five at 5mg of RG7203 when compared to all other 1768 connections of cerebrum structures.

## Discussion

Here we tested the relationship between changes in apathy and striatal connectivity during treatment with the PDE10 inhibitor RG7203 which was given as add-on to antipsychotic medication. Compared to placebo, treatment with neither 5mg nor 15mg of RG7203 resulted in significant changes in apathy or striatal connectivity. However, the effect of RG7203 on apathy showed clinically relevant variation that was reflected in striatal connectivity. With 5mg RG7203 vs. placebo, increased connectivity between vCaud and PCG as well as between vCaud and ACC was associated with apathy improvement and vice versa. A similar association was observed for 15 mg of RG7203 although it did not reach statistical significance. Importantly, such correlations were not observed for the negative symptom dimension of diminished expression.

The relationship between changes in apathy and vCaud-PCG/ACC connectivity supports the association of these cortico-striatal components with apathy [8] and aligns with current theories about the functions of vCaud, ACC and PCG. The network of vCaud, ACC and PCG has been implicated in integration of cognitive and reward processing, and impairment in these domains is assumed to be a primary driver of apathy [8, 30-36]. A recent study proposed that parts of ACC and ventral striatum play a key role in the generation of apathy across neurological disorders [37]. A foregrounded relevance of ventral striatum and ACC for apathy would be in line with our finding that changes in apathy and vCaud-ACC are correlated but may also explain why we did not find a relationship between changes in apathy and connectivity for most of the other tested connectivity pairs. The only other connectivity with a significant relationship to apathy changes pertained the connectivity between vCaud and PCG. Interestingly, ACC and PCG are adjacent structures in the medial part of the brain. Apart from that, correlations between changes in functional connectivity and apathy rarely showed high magnitudes in our cerebrum-wide analysis. This aligns with recent theories that view schizophrenia as a disorder of discrete neural circuits rather than a diffuse brain dysfunction [23, 38, 39].

We also observed that lower vCaud-PCG/ACC connectivity during placebo treatment was associated with larger improvement of apathy during treatment with RG7203. This is in line with our main finding that increased vCaud-PCG/ACC connectivity is a correlate of improved apathy with RG7203 vs. placebo. However, an interesting hypothesis could also be that lower vCaud-PCG/ACC connectivity increases the probability that patients benefit from RG7203 (with the caveat that the placebo condition is not the same as a baseline condition). The relationship between changes in apathy with RG7203 and striatal connectivity at baseline could be investigated in larger clinical studies that are designed for this purpose. If resting-state striatal connectivity at baseline would inform the benefit patients can expect from RG7203, this would advance biologically informed treatment options for apathy and pave the way for more personalized treatments in schizophrenia.

There are several limitations to our study. First, we cannot rule out the possibility that apathy changes with RG7203 vs. placebo simply reflect longitudinal apathy fluctuations with no relation to RG7203. The extent of interindividual differences in apathy modulation and the similar apathy response to both doses of RG7203 provide arguments against this possibility. Considering the limited test-retest reliability of functional connectivity [40, 41], it is also possible that the observed changes in functional connectivity with RG7203 vs. placebo reflect longitudinal connectivity fluctuations with no relation to RG7203. The observed consistency in connectivity alterations across both doses of RG7203, especially for the connectivity pairs vCaud-PCG and vCaud-ACC, may at least indicate that this is less probable. Moreover, the treatment duration was relatively short, and it is unknown whether we would have found significantly altered apathy and striatal connectivity after a longer treatment course with RG7203. The sample size was underpowered for detecting smaller effects. Furthermore, the complexity of brain circuitry and its interaction with pharmacological agents calls for a cautious interpretation of the observed correlations. Future studies with larger sample sizes, longer treatment durations, and comprehensive assessments of striatal connectivity are required to substantiate and expand upon these findings. In conclusion, our study elucidated neurobiological underpinnings for the apathy changes under RG7203. The observed relationships between vCaud-PCG/ACC connectivity and apathy changes opens avenues for further research and personalized therapy.

## Data Availability

All data produced in the present study are available upon reasonable request to the authors.

## Acknowledgements

We thank Markus Abt, Paul Tamburri, Christopher Chatham, Michael J. Frank, Anne G.E. Collins, David P. Walling, Rick Mofsen, Daniel Gruener, Lev Gertsik, Jeff Sevigny and Sanjay Keswani for their substantial contributions to the project^4^.

## Author contributions

W. O. co-conceptualized the data analysis, co-analyzed and interpreted the data, performed the statistical analysis, wrote and refined the manuscript. D.U. conceived, designed, implemented and supervised the original study together with collaborators and contributed to the manuscript. G.C. and G.-Y. H. supported data analysis. G.G. and Š.H. provided the data set. Š.H. and J.D. performed the MRI setup at all study centers, including the MRI sequence setup, operators’ training, and the MRI-specific study documentation; and additionally implemented the MRI study in the centers including QC reviews and data transfers and data curation. M.K. and W.S. reviewed and edited the manuscript. P. H. initiated and co-conceptualized the data analysis, co-interpreted and supervised the study, and refined the manuscript. All authors approved and refined the final version of the manuscript.

