## SupplementaryMaterial for "Relationship between Striatal Connectivity and Apathy during Phosphodiesterase 10 Inhibition in Schizophrenia"

**Supplementary Material**

**I. Supplementary figures:**

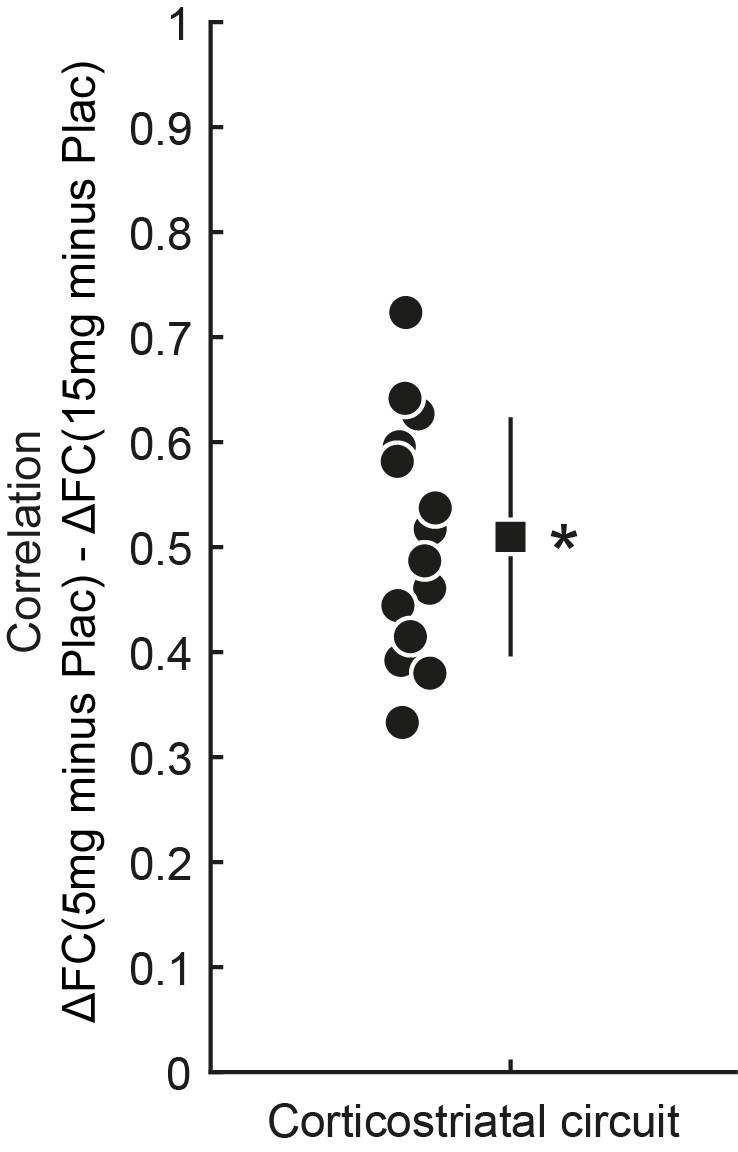

**Supplementary Fig. 1: Correlation between striatal connectivity alterations at 5mg and 15mg RG7203.** For all connectivity pairs shown in Fig. 1b, the correlation between connectivity alterations at 5mg and 15mg RG7203 versus placebo is illustrated. Asterisk indicates p<0.05 (Wilcoxon signed-rank test, n = 14 correlations).

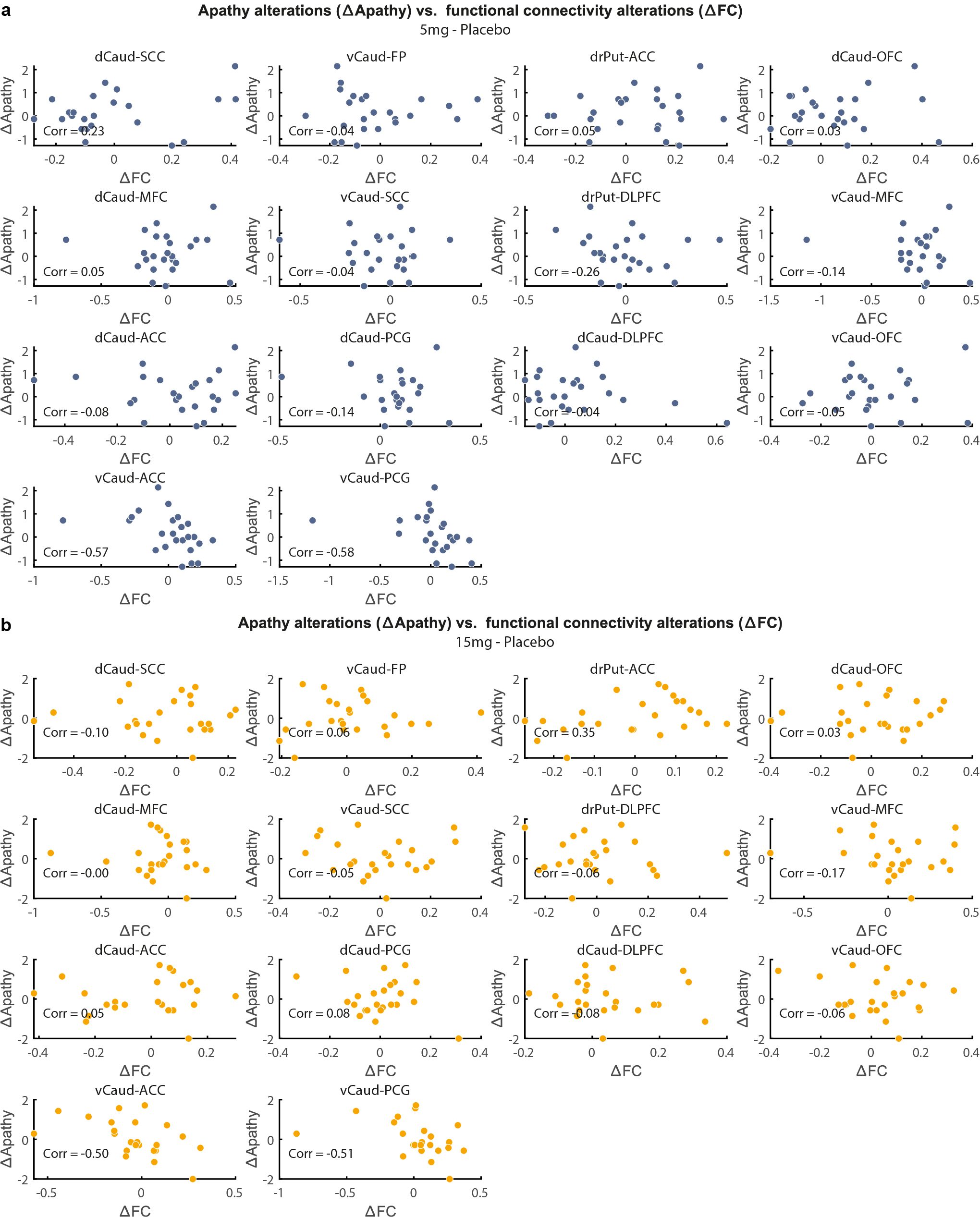

**Supplementary Figure 2: Alterations in apathy vs. alterations in striatal connectivity with RG7203 vs. placebo. a.** Individual values for ∆Apathy and ∆FC are shown for 5mg RG7203 vs. placebo. The Spearman correlation is shown as text inlay for each connectivity pair. **b.** Same for 15mg RG7203 vs. placebo.

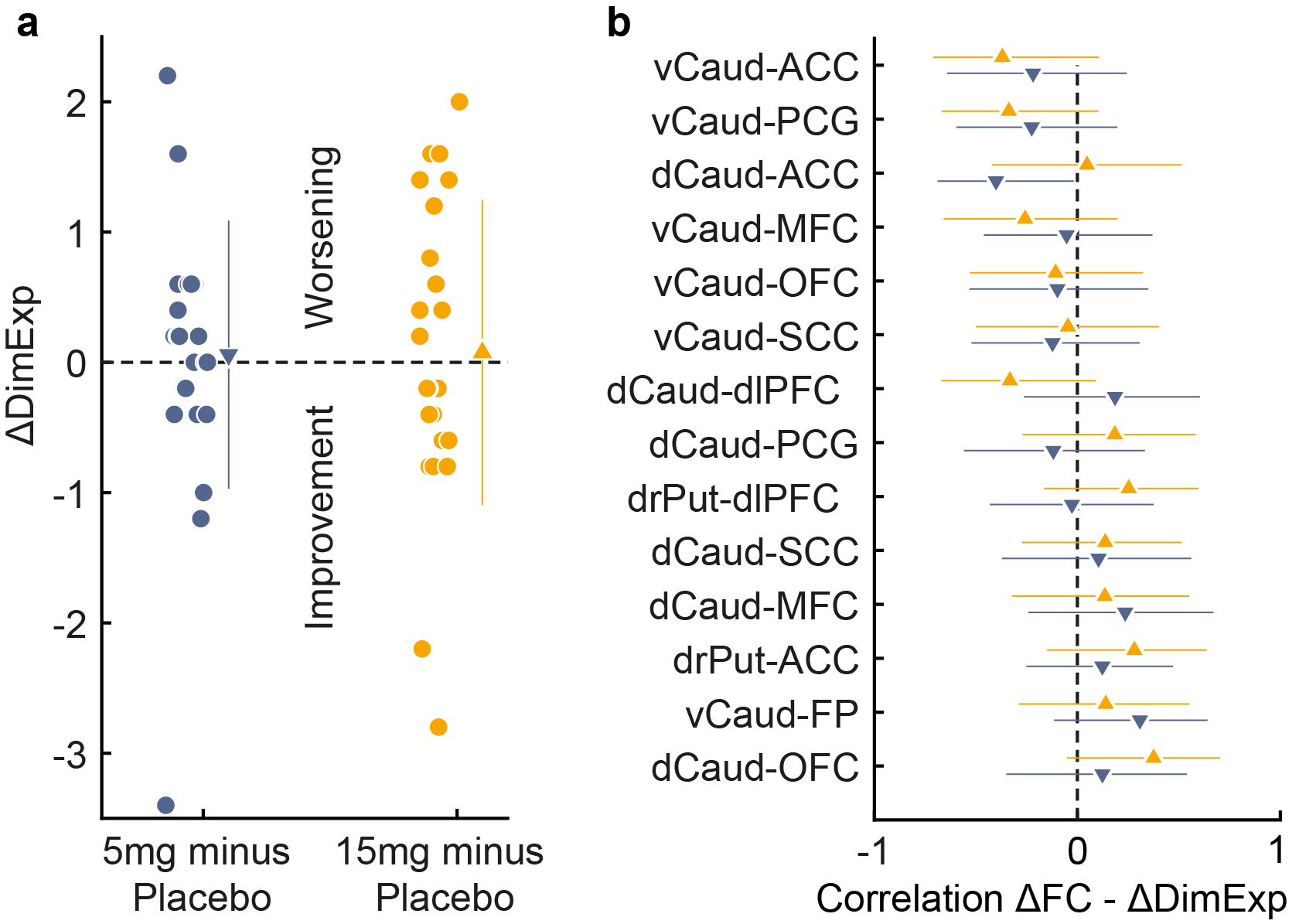

**Supplementary Figure 3: Correlation between alterations in diminished expression and striatal connectivity. a.** Alterations in diminished expression (∆DimExp) when 5mg or 15mg RG7203 are given instead of placebo (5mg minus placebo and 15mg minus placebo); filled circles represent individual subjects; triangles with error bars show means and standard deviations. **b.** Correlation between ∆DimExp and ∆FC is illustrated for the investigated corticostriatal circuit. The error bars correspond to the 95% confidence interval.

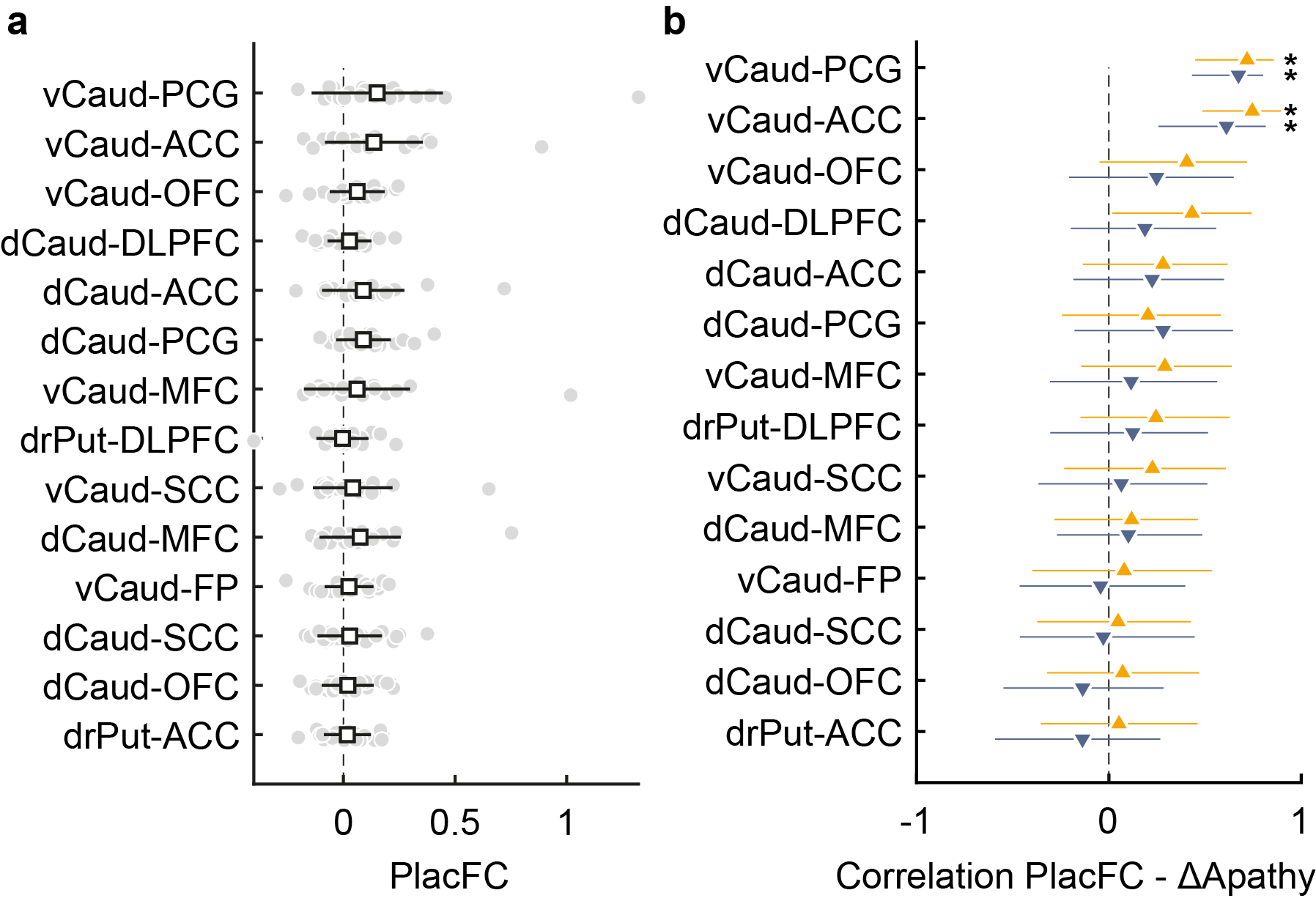

**Supplementary Figure 4: Correlation between striatal connectivity under placebo and apathy alterations with 5mg or 15mg of RG7203 versus placebo. a.** Functional connectivity of the cortico-striatal circuit under placebo; filled circles represent individual subjects; squares with error bars show means and standard deviations **b.** Correlation between functional connectivity under placebo and ∆Apathy is illustrated for the cortico-striatal circuit. The error bars correspond to the 95% confidence interval. Asterisks indicate p<0.05.

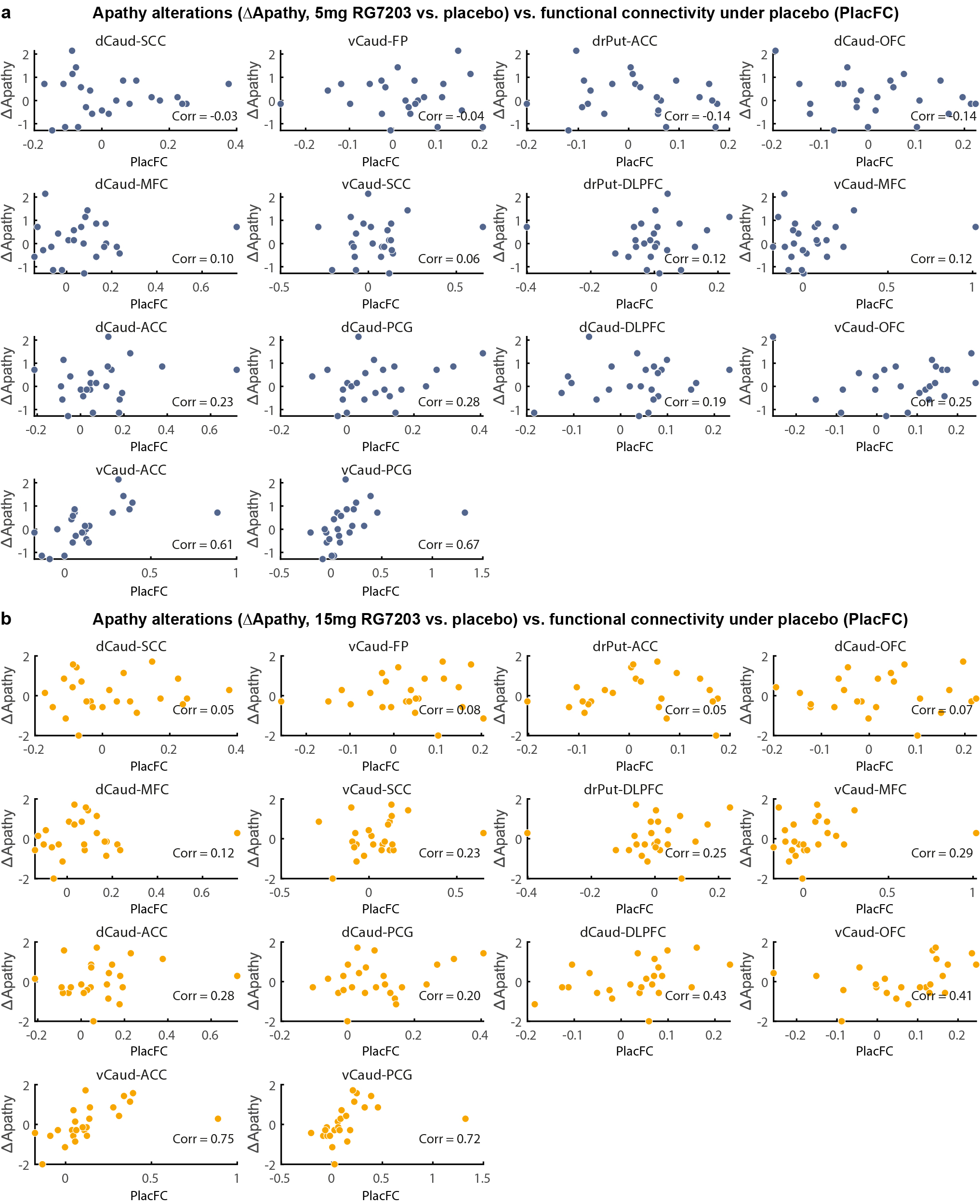

**Supplementary Figure 5: Alterations in apathy with RG7203 vs. placebo and striatal connectivity under placebo. a.** Individual values for ∆Apathy (5mg RG7203 minus placebo) and functional connectivity under placebo (PlacFC) are shown. The Spearman correlation is shown as text inlay for each connectivity pair. **b.** Individual values for ∆Apathy (15mg RG7203 minus placebo) and functional connectivity under placebo (PlacFC) are shown.

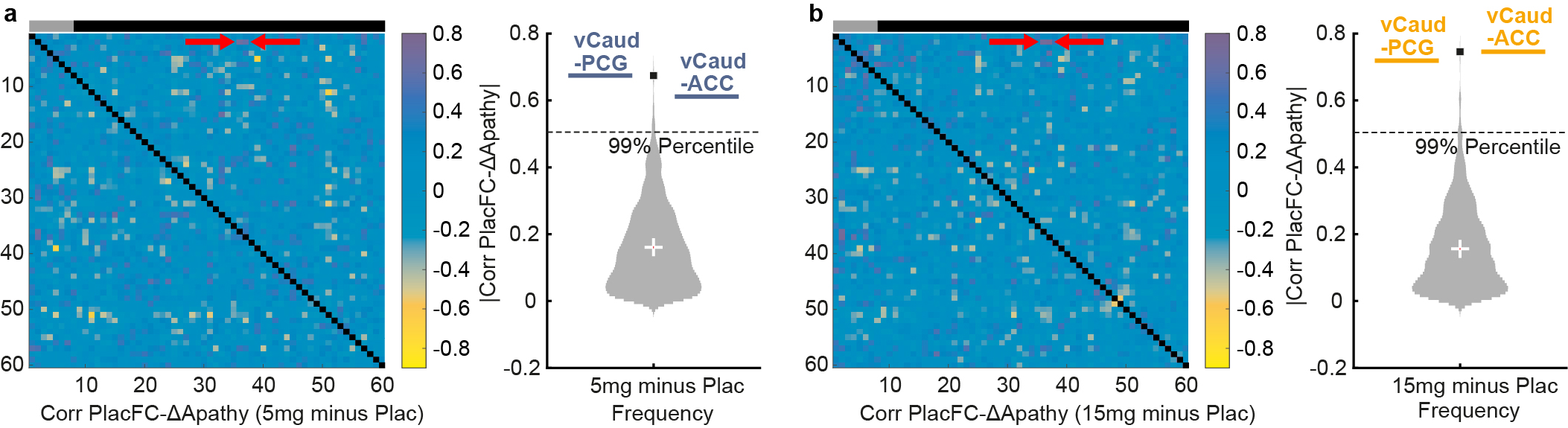

**Supplementary Figure 6: Relationship between apathy changes with RG7203 vs. placebo and connectivity under placebo conditions for all combinations of cerebrum structures.**

**a.** 5mg RG7203 compared to placebo. Left panel: Functional connectivity under placebo conditions (PlacFC) has been computed for all possible combinations of the 60 anatomical cerebrum structures and correlated with apathy changes (∆Apathy) at 5mg RG7203; gray bar indicates striatal regions, black bar extrastriatal regions; correlation of ∆Apathy and placebo connectivity between vCaud and PCG/ACC is indicated by red arrows. Right panel: Violin plot showing the absolute value of all 1770 unique correlations between PlacFC and ∆Apathy; correlation of ∆Apathy and placebo connectivity between vCaud and PCG/ACC is indicated by blue bars. The black square indicates the maximum of all correlations. **b.** 15mg RG7203 vs. placebo: Same computations and conventions as in a, but correlation between ∆Apathy and placebo connectivity for vCaud and PCG/ACC is indicated by orange instead of blue bars in the right panel.

**II. Eligibility criteria[16]:**

**Inclusion Criteria:**

Patients must meet the following criteria for study entry:

1. A Diagnostic and Statistical Manual of Mental Disorders (DSM)-5 diagnosis of schizophrenia as established by the Structured Clinical Interview for DSM Clinical Trials version (SCID-5-CT) at screening.

2. Age 18–50 years (inclusive).

3. Outpatient with no hospitalization for worsening of schizophrenia within 3 months prior to screening (hospitalization for social management within this time was acceptable).

4. Medically stable over 1 month and psychiatrically stable without symptom exacerbation over the previous 3 months prior to screening.

5. Males and females with no childbearing capacity; females must be either surgically sterile (by means of hysterectomy, bilateral oophorectomy or tubal ligation) or post-menopausal for at least 1 year (confirmed by follicle stimulating hormone and estradiol, if not on hormone replacement).

6. Has a caregiver or other identified responsible person considered reliable by the investigator to provide support to the patient to help ensure compliance with study treatment, study visits and protocol procedures, and who preferably is also able to provide input helpful for completing study rating scales.

7. Body mass index > 18.5 kg/m 2 and < 35 kg/m2.

8. Fluent in English, even if English is not the primary language.

9. Able and willing to provide written informed consent according to International Council for Harmonisation of Technical Requirements for Pharmaceuticals for Human Use and local regulations and to comply with the study protocol. (If the patient has a legal representative, the informed consent must be signed by this person as well.)

10. Able to complete study procedures.

*Symptom severity at screening:*

11. The patient has a Clinical Global Impression-Severity scale (CGI-S) score ≥ 3 (mildly ill).

12. The patient has a score of < 4 (moderate) on the following Positive and Negative Syndrome Scale (PANSS) items:

a) P7 (hostility)

b) G8 (uncooperativeness)

13. Total score of ≥ 18 on the PANSS negative symptom factor score (items scored

1–7 for a maximum possible score of 49).

14. PANSS item G6, depression score of ≤ 4 (moderate).

15. Depressive symptoms, defined as a score of ≤ 8 on the Calgary Depression

Rating Scale for schizophrenia.

*Antipsychotic treatment:*

16. On stable treatment, that is 6 weeks without change, with no more than 2 antipsychotics (oral and long-acting injectable formulations of the same medication are considered to be two different antipsychotics) prior to screening. Antipsychotic regimen:

c) Patients must be on a “primary” antipsychotic and may be on a secondary antipsychotic. The amount of the secondary antipsychotic has to be equal or less than the equivalent dose of the primary antipsychotic and the sum of the primary and secondary antipsychotics has to be ≤ 6 mg of risperidone equivalents.

d) The allowed “primary” antipsychotics are quetiapine, paliperidone, risperidone, aripiprazole, lurasidone and ziprasidone. Antipsychotics have to be used in the dose range according to the prescribing information approved in the US. In the case that the primary antipsychotic is a first-generation antipsychotic (oral or injectable) an exception may be granted if discussed and clearly documented between the investigator and the Sponsor (Translational Medicine Leader or delegate) and approved by the Sponsor.

**Exclusion Criteria:**

Patients who meet any of the following criteria will be excluded from study entry:

1. Moderate to severe substance use disorder within 6 months (excluding nicotine or caffeine) as defined by DSM-5.

2. Positive urine drug screen for amphetamines, methamphetamines, opiates, buprenorphine, methadone, cannabinoids, cocaine and barbiturates.

3. Other current Axis I diagnosis requiring exclusion (e.g., bipolar disorder, schizoaffective disorder, major depressive disorder) based on DSM-5.

4. The patient is at significant risk of suicide or harming him- or herself or others according to the Investigator’s judgment.

5. History of neuroleptic malignant syndrome.

6. A prior or current general medical condition that might be impairing cognition or other psychiatric functioning (e.g., migraine headaches requiring prophylaxis treatment, head trauma, dementia, seizure disorder, stroke, neurodegenerative, inflammatory, infectious, neoplastic, toxic, metabolic, endocrine etc.).

7. A movement disorder due to antipsychotic treatment not currently controlled with anti-EPS (extrapyramidal symptoms) treatment or another movement disorder which might affect the ratings on the EPS scales (e.g., Parkinson’s disease).

8. The patient has a score > 2 (mild) in any of the four CGI-S items of the Extrapyramidal Symptom Rating Scale – Abbreviated; Parkinsonism, akathisia, dystonia, and tardive dyskinesia at screening or on Day -1 and was requiring anti-Parkinson medication including anticholinergic drugs.

9. History of HIV infection, or history of Hepatitis B infection within the previous year, or history of Hepatitis C infection which had not been adequately treated.

10. QTcF interval > 450 msec (470 msec for females) or other clinically significant abnormality on screening electrocardiogram based on centralized reading.

11. Clinically significant abnormalities in laboratory safety test results (including hepatic and renal panels, complete blood count, chemistry panel and urinalysis). In case of uncertain or questionable results, tests performed during screening may be repeated before randomization to confirm eligibility or may be accepted if they are, in the opinion of the Investigator, clinically insignificant.

10. Significant or unstable physical condition which in the Investigator’s judgment might require a change in medication or hospitalization within the next 3 months.

11. Receipt of an investigational drug within 90 days or 5 times the half-life of the investigational drug, whatever is longer, prior to screening.

12. Previously received RO5545965.

13. Electroconvulsive treatment within 6 months prior to screening.

14. Currently on treatment with olanzapine or clozapine.

15. Having received olanzapine or clozapine within 3 months of screening.

16. On more than one antidepressant, or if on one antidepressant, a change in dose within 4 weeks prior to screening.

17. Change in benzodiazepine or sleep medication regimen within 2 weeks (regimen could be as needed or continuous treatment) prior to screening.

18. Change in anti-EPS medication within two weeks prior to screening.

19. Use of prohibited medications taken within 14 days or within 5 times the elimination half-life of the medication before the first study drug administration (whichever is longer), except for allowed medication:

• Concomitant therapy includes any medication, e.g., prescription drugs, over the counter drugs, approved dietary and herbal supplements, nutritional supplements and any nonmedication interventions (e.g., individual psychotherapy, cognitive behavioral therapy, smoking cessation therapy, and rehabilitative therapy) used by a patient from 6 weeks prior to screening until the follow-up visit.

20. Use of any strong or moderate inhibitor of CYP3A or CYP2C8 and any inducer of CYP3A within 14 days or within 5 times the elimination half-life of the medication (whichever is longer) before the first study drug administration, including but not limited to the following: rifampicin, carbamazepine, St. John's wort, ketoconazole, itraconazole, fluconazole, erythromycin, cimetidine, fluoxetine, gemfibrozil and phenytoin.

21. Use of any other nutrients known to modulate CYP3A activity (e.g., grapefruit containing products) within 1 week before the first study drug administration.

22. The patient is a member of the study personnel or of their immediate families or is a subordinate (or immediate family member of a subordinate) to any of the study personnel.
